# Digital Photo-Elicited Storytelling and Alpha-Band EEG Dynamics in Older Adult–Caregiver Dyads

**DOI:** 10.64898/2026.07.09.26357346

**Authors:** Supalak Khemthong, Winai Chatthong

## Abstract

Digital technologies can support meaningful social interaction by providing relevant prompts for memory and shared reflection. Mobile-phone photography offers an accessible medium for older adults and caregivers to construct stories and engage relationally. However, psychophysiological evidence on digital photo-supported storytelling in older adult–caregiver dyads remains limited. This study examined alpha-band EEG dynamics during photo-elicited storytelling in two museum settings.

Thirty-two older adult–caregiver dyads used mobile-phone photography to capture meaningful scenes, selecting one photo as a prompt for naturalistic storytelling. EEG was recorded during eyes-closed resting, eyes-open resting, storytelling, and listening. We analyzed task-related relative alpha power using a 10-electrode set. We also tested associations between Cz alpha power and MoCA scores, and explored dyadic alpha-band inter-brain similarity using within-site shuffled-dyad surrogate comparisons.

Alpha power was higher during eyes-closed resting and lower during storytelling and listening relative to eyes-open resting, indicating task-related modulation. Associations between MoCA scores and Cz alpha power were weak and did not survive false discovery rate correction. Dyad-level inter-brain similarity during storytelling was modestly higher than shuffled-dyad estimates, but nonsignificant after correction across conditions.

These findings suggest photo-elicited storytelling is a meaningful medium for studying cognitive-social engagement in dyads. Alpha-band EEG was sensitive to narrative interactions, although cognition and dyadic similarity effects were modest. This study contributes to technology-supported human behavior research by demonstrating how digital image prompts structure naturalistic interaction while enabling real-world psychophysiological measurement.

## 1. Introduction

Digital technologies increasingly shape how people remember, communicate, and participate in social life. In later life, simple and familiar tools such as mobile-phone photography can support autobiographical reflection, emotional expression, and interpersonal communication. Photographs are not merely records of experience; they can become prompts through which people organize memories, construct meaning, and communicate with others. Digital photo-elicitation may therefore provide a useful medium for studying human behavior in naturalistic social contexts.

Older adults’ cognitive and social engagement is shaped not only by individual cognitive status but also by meaningful activity and interpersonal participation. Socially integrated and cognitively stimulating activities have been associated with reduced dementia risk or slower cognitive decline (Fratiglioni et al., 2004; Livingston et al., 2020). However, the mechanisms through which technology-supported activities engage attention, memory, communication, and interpersonal coordination remain incompletely understood. This gap is important because real-world activities such as photo-based storytelling require autobiographical memory retrieval, semantic organization, affective expression, and partner-directed communication.

Digital photo-elicited storytelling provides a structured but naturalistic way to examine these processes. Unlike standardized laboratory stimuli, personally selected photographs allow participants to construct narratives around meaningful objects, places, and experiences. At the same time, the use of a common digital medium provides sufficient procedural structure for research measurement. In older adult–caregiver dyads, this approach may support interaction by giving both partners a shared visual reference while allowing narrative content to remain personally meaningful.

Caregivers play an important role in this interaction. In storytelling activities, caregivers are not only companions or facilitators; they listen, respond, interpret, and help sustain shared meaning. Listening in this context is not passive hearing. It involves attention, emotional presence, patience, openness, and engagement with another person’s memories and meanings. Digital photo-elicited storytelling may therefore provide a practical medium through which caregivers and older adults can engage in relational communication, memory sharing, and mutual reflection.

Electroencephalography provides a non-invasive method for examining physiological brain activity during such technology-supported interaction. Alpha-band activity is particularly relevant because it is sensitive to attentional control, access to stored information, and task-related engagement (Klimesch, 2012; Pfurtscheller & Lopes da Silva, 1999). In aging research, altered alpha and alpha/beta dynamics have been linked to memory and cognitive performance (Griffiths et al., 2021; Healey & Kahana, 2020; Karlsson & Sander, 2022). Alpha-band EEG activity may therefore provide a useful sensor-level index for examining cognitive engagement during digital photo-supported storytelling and listening.

Dyadic interaction adds another layer of complexity because cognitive processing unfolds across interacting individuals. Hyperscanning and relational neuroscience studies suggest that inter-brain measures may be associated with shared attention, interpersonal coordination, social connection, and cooperative behavior (De Felice et al., 2025; Dumas et al., 2010; Hinvest et al., 2025; Kinreich et al., 2017; Wolff & Dumas, 2026). However, similarity between partners can also arise from shared sensory input, common task structure, turn-taking, or environmental context. Therefore, dyad-level EEG similarity should be interpreted cautiously and compared with surrogate or shuffled-dyad estimates when possible.

The present study examined alpha-band EEG dynamics during digital photo-elicited storytelling in older adult–caregiver dyads. Participants used mobile-phone photography during a museum visit to capture personally meaningful objects, scenes, or exhibition spaces. These digital photographs then served as prompts for structured but naturalistic storytelling and listening. We examined whether alpha power was modulated during storytelling and listening relative to eyes-open resting, whether Cz alpha power was associated with cognitive performance measured using MoCA scores, and whether true dyads showed greater alpha-band spatial similarity than within-site shuffled dyads. By combining digital photo-elicitation, naturalistic storytelling, and EEG measurement, this study aimed to characterize cognitive and social engagement during technology-supported human interaction in later life.

## 2. Methods

### 2.1. Study design and setting

This study used a naturalistic, within-participant, dyadic design to examine digital photo-elicited storytelling in older adult–caregiver dyads. The study was conducted in two museum settings in XXX: Museum XXX and the XXX Museum. These settings were selected because they provided culturally meaningful objects, images, exhibition spaces, and narratives that could support autobiographical reflection and social communication.

The study combined a technology-supported storytelling task with EEG recording. Mobile-phone photography was used as the digital medium through which participants selected personally meaningful museum experiences and transformed them into storytelling prompts. EEG was used as supplementary psychophysiological evidence to examine alpha-band activity during resting, storytelling, and listening conditions. The study was not designed to evaluate a clinical intervention or to diagnose cognitive or emotional states.

### 2.2. Participants

Thirty-two older adult–caregiver dyads participated in the study, comprising 64 participants. Sixteen dyads were recruited from Museum XXX and 16 dyads were recruited from the XXX Museum. Older adult participants were aged 60 years or older and attended the activity with a caregiver, family member, paid caregiver, or volunteer companion. Caregivers were aged 25–59 years.

Participants were recruited through community outreach and museum education activities. All participants provided written informed consent before participation. They were informed about the voluntary nature of the study, confidentiality of their data, and compensation for participation. The study received ethical approval from the relevant institutional review board.

Participant characteristics are reported in Table 1, including participant role, museum site, age, sex, cognitive screening scores, depressive symptom scores, and other demographic variables where available.

### 2.3. Inclusion and exclusion criteria

Older adults were eligible if they were aged 60 years or older, legally competent, able to communicate in Thai, and able to use a mobile phone to take photographs during the museum activity. They were screened using the XXX version of the Patient Health Questionnaire-9 and were included if their scores indicated no to mild depressive symptoms. Cognitive status was screened using the XXX version of the Montreal Cognitive Assessment, and participants scoring above 16 were eligible, indicating cognitive performance ranging from normal cognition to possible mild cognitive impairment.

Caregivers were eligible if they were aged 25–59 years, legally competent, able to communicate in XXX, able to use a mobile phone to take photographs, and accompanying the older adult as a relative, family member, paid caregiver, or volunteer companion.

Participants were excluded if they had neurological disorders associated with severe cognitive impairment, including dementia, stroke, Parkinson’s disease, or delirium; moderate-to-severe psychiatric illness; active hallucinations; current medication or treatment known to substantially influence EEG recordings; or abnormal involuntary movements that could interfere with EEG acquisition.

### 2.4. Digital photo-elicited storytelling task

The storytelling task used a digital photo-elicitation approach. Before EEG recording, each dyad visited six exhibition rooms in the museum for approximately 60 minutes. During the visit, older adults and caregivers used mobile phones to capture photographs of museum objects, scenes, or exhibition spaces that they found personally meaningful, memorable, emotionally salient, or relevant to later-life well-being. Participants could take photographs independently or take photographs for one another.

After the museum visit, each participant selected one photograph that was most personally meaningful. This photograph served as a digital prompt for the storytelling interaction. Participants were asked to prepare a brief narrative guided by three standardized questions: (1) What is shown in the photograph? (2) Why was this photograph meaningful or impressive? and (3) How does this image relate to health, well-being, or happiness in later life?

The task was designed to balance ecological validity with procedural consistency. Narrative content was not identical across dyads because the study aimed to capture personally meaningful storytelling. However, the digital-photo prompt, storytelling sequence, task instructions, EEG timing, and recording procedures were standardized across dyads and museum sites.

### 2.5. Caregiver listening and dyadic interaction roles

The storytelling protocol included both speaking and listening roles. During the EEG-recorded interaction, the older adult first narrated the selected photograph while the caregiver listened. The caregiver was allowed to respond naturally, ask clarifying questions, or share related reflections without leading the narrative. The caregiver then became the storyteller, and the older adult became the listener.

In this study, caregiver listening was treated as an active component of digital photo-elicited storytelling rather than a passive background condition. Listening in this context involved attention to the older adult’s story, emotional presence, openness to another person’s memories and meanings, and engagement with shared reflection. However, the study did not claim to directly measure empathy as a psychological construct. Instead, EEG activity during listening was interpreted as sensor-level physiological activity occurring during a task context designed to elicit attentive and relational engagement.

### 2.6. Experimental procedure

The experimental procedure consisted of six sequential phases: participant enrolment, museum visit, photograph selection, EEG recording, post-recording care, and data analysis.

First, participants completed enrolment. Demographic and background information was collected, including age, sex, education, socioeconomic background, health history, caregiver relationship, and museum visit history. Cognitive and psychological screening was conducted using the MoCA and PHQ-9. Written informed consent was obtained from all participants.

Second, each dyad visited six exhibition rooms in the museum for approximately 60 minutes and used mobile-phone photography to capture personally meaningful museum objects, scenes, or exhibition spaces.

Third, after the museum visit, each participant selected one photograph to serve as the digital prompt for the storytelling interaction.

Fourth, EEG recording was conducted while participants wore the EEG cap. EEG setup and signal preparation lasted approximately 11 minutes. EEG data were recorded during resting and interaction conditions. Eyes-closed resting EEG was recorded for 3 minutes, followed by eyes-open resting EEG for 6 minutes while dyad members sat facing one another without storytelling.

The storytelling interaction was then recorded for 10 minutes, consisting of 5 minutes in which the older adult narrated the selected photograph while the caregiver listened, followed by 5 minutes in which the caregiver narrated while the older adult listened.

Fifth, after EEG recording, conductive gel was removed from the hair and scalp using dry and wet tissues to ensure participant comfort.

Finally, EEG data were processed for spectral analysis and alpha-band inter-brain similarity analysis. Digital photographs and narrative materials were used to contextualize the storytelling interaction but were not treated as fixed experimental stimuli.

### 2.7. Cognitive and psychological measures

Cognitive performance was assessed using the XXX version of the Montreal Cognitive Assessment. The MoCA evaluates visuospatial and executive function, attention, memory, language, abstraction, and orientation. Depressive symptoms were assessed using the XXX version of the Patient Health Questionnaire-9, a nine-item self-report measure scored from 0 to 27. These measures were used for screening and participant characterization rather than for clinical diagnosis. MoCA scores were also used as a continuous index of cognitive performance in correlation analyses.

### 2.8. EEG recording and preprocessing

EEG signals were recorded using a BrainMaster Discovery 24E QEEG system (K150498; BrainMaster Technologies, Inc.). The system included a 19-position electrode cap arranged according to the international 10–20 system and was connected to a 12-channel signal acquisition unit. Recordings were obtained during eyes-closed resting, eyes-open resting, storytelling, and listening conditions using the same hardware and montage settings.

Before recording, electrode impedance was checked and maintained below 5 kΩ for all channels, with inter-electrode impedance differences kept within 2 kΩ. EEG data were sampled at 250 Hz and referenced to linked ears. Continuous EEG signals were notch filtered at 50 Hz to reduce line-noise contamination and bandpass filtered from 1 to 30 Hz before spectral analysis.

EEG preprocessing and spectral analysis were conducted offline using NeuroGuide Deluxe QEEG software, version 3.0.7.3. Automated artifact detection was applied using an amplitude threshold of ±100 µV to exclude segments containing gross artifacts, including eye blinks, swallowing, chewing, head movement, and body movement. Artifact sensitivity was set to high to reduce contamination from drowsiness-related activity and eye movements. Amplitude selection was adjusted using the multiplier method, with values set between 0.9 and 1.0.

Independent component analysis was applied to identify and remove components associated with ocular and muscle artifacts. Automated preprocessing was followed by manual visual inspection to ensure that retained segments were suitable for spectral analysis and topographical mapping. A minimum of 60 seconds of artifact-free EEG data was retained for each analyzed condition.

Power spectral density was estimated using Welch’s method with 2-second epochs and 50% overlap. NeuroGuide generated standard spectral estimates across delta, theta, alpha, and beta frequency bands. In the inferential analysis, alpha-band activity was specified as the primary frequency band of interest to reduce the number of statistical comparisons and to focus on cognition-related oscillatory activity during digital photo-supported storytelling and listening. Relative alpha power was calculated as alpha power divided by total power across the 1–30 Hz range.

Topographical maps were generated as descriptive visualizations of sensor-level alpha-power patterns across resting, storytelling, and listening conditions. Z-score brain maps derived from the NeuroGuide normative database were used descriptively and were not interpreted as clinical diagnostic maps or evidence of localized neural generators. Individual alpha peak frequency was not controlled for in the present analysis.

### 2.9. Alpha-band inter-brain similarity analysis

Alpha-band inter-brain similarity was defined as dyad-level similarity in sensor-level alpha-power patterns between an older adult and their caregiver during the same experimental condition. This term was used instead of stronger claims about neural synchrony because the present analysis was based on spatial similarity in spectral power rather than continuous time-resolved hyperscanning connectivity.

For each dyad and condition, relative alpha power values were extracted from a predefined 10-electrode sensor-level set: Fp1, Fp2, F3, F4, F8, T3, T4, Fz, Cz, and Pz. These values were arranged as spatial alpha-power patterns separately for the older adult and caregiver. Inter-brain similarity was calculated using Pearson’s spatial correlation between the older adult’s and caregiver’s 10-electrode alpha-power patterns within each dyad. Correlation coefficients were transformed using Fisher’s z for statistical reporting. Higher Fisher’s z values indicated greater similarity in alpha-power distribution between dyad members.

To address the possibility that similarity was driven by shared task structure, common sensory input, museum environment, or procedural artifacts, a surrogate-dyad robustness check was conducted. Older adults and caregivers were randomly re-paired within the same museum site, thereby preserving site-specific museum context while disrupting the original dyadic pairing. Alpha-band inter-brain similarity was recalculated for each shuffled pairing across 10,000 permutations to generate a surrogate distribution of chance-level dyadic similarity. Observed dyad-level similarity was then compared with the surrogate distribution to determine whether true dyads showed greater alpha-band spatial similarity than would be expected from participants exposed to broadly similar museum and task contexts but not interacting as a dyad. The resulting permutation p value was calculated as the proportion of surrogate similarity values equal to or greater than the observed similarity value.

### 2.10. Statistical analysis

Statistical analyses were conducted using IBM SPSS Statistics. Participant characteristics were summarized using means and standard deviations for continuous variables and counts and percentages for categorical variables.

Alpha-band activity was specified as the primary frequency band of interest. To reduce the number of statistical comparisons and avoid post hoc interpretation of individual electrode findings, inferential EEG analyses were restricted to the predefined 10-electrode sensor-level alpha set. This set provided coverage of frontal, frontotemporal, temporal, central, and parietal midline regions relevant to attentional engagement, narrative processing, and social-cognitive task participation.

To evaluate task-related alpha modulation during digital photo-elicited storytelling and listening, alpha power was analyzed across four conditions: eyes-closed resting, eyes-open resting, storytelling, and listening. The eyes-open resting condition was used as the within-participant baseline to reduce inter-individual variability and to control for visual activation effects. For each participant, task-related difference scores were calculated as:

Δ Alpha Power = Relative Alpha PowerCondition − Relative Alpha PowerEyes Open

Difference scores were summarized as mean differences, standard deviations, and 95% confidence intervals by participant role and condition.

Before inferential analyses, normality of Δ Alpha Power values was assessed using the Shapiro–Wilk test. When Δ Alpha Power values were normally distributed, paired-samples t-tests were used to examine within-participant differences between eyes-open resting and eyes-closed resting, storytelling, or listening conditions. When normality assumptions were not met, Wilcoxon signed-rank tests were used. Effect sizes were reported using Cohen’s dz for paired-samples t-tests and rank-based effect sizes for nonparametric comparisons where appropriate.

To address multiple comparisons, p values from planned condition-based comparisons were adjusted using the Benjamini–Hochberg false discovery rate procedure. Both uncorrected p values and FDR-adjusted q values were reported. Findings that remained significant after FDR correction were interpreted as statistically robust within the exploratory framework, whereas findings that did not survive correction were interpreted cautiously as hypothesis-generating.

Associations between alpha power and cognitive measures were examined using correlation analyses. Cz alpha power was retained as a focused central region-of-interest analysis for cognition-related associations because central alpha activity was considered theoretically relevant to attentional engagement and sensorimotor integration during storytelling and listening. Normality of the relevant EEG and behavioral variables was assessed using the Shapiro–Wilk test before correlation analysis. Pearson’s correlation coefficients were used when both variables were normally distributed, whereas Spearman’s rank correlation coefficients were used when at least one variable was not normally distributed. Correlations were examined between relative alpha power and MoCA scores. Correlation coefficients, p values, FDR-adjusted q values, and 95% confidence intervals were reported.

Alpha-band inter-brain similarity analyses were conducted using Fisher’s z-transformed Pearson spatial correlations derived from the 10-electrode alpha-power patterns. Within-site surrogate-dyad analyses were used to evaluate whether observed dyad-level similarity exceeded chance pairing while preserving museum-site context. Permutation p values were reported, and FDR correction was applied across condition-specific surrogate comparisons.

A post hoc sensitivity analysis was conducted using G*Power 3.1 to estimate the minimum detectable effect size for participant-level and dyad-level analyses. Given the modest sample size and exploratory dyadic design, small effects were interpreted cautiously.

## 3. Results

### 3.1. Participant characteristics

Thirty-two older adult–caregiver dyads participated in the digital photo-elicited storytelling protocol, comprising 64 participants in total. The sample included 32 older adults and 32 caregivers recruited from two museum settings: Museum XXX and the XXX Museum, with 16 dyads from each site.

As shown in Table 1, the mean age of the total sample was 56.70 years (SD = 14.92). Older adults had a mean age of 69.38 years (SD = 5.68), while caregivers had a mean age of 44.03 years (SD = 9.43). The sample was predominantly female, with 49 female participants (76.6%) and 15 male participants (23.4%). Mean MoCA scores were 24.53 (SD = 3.95) among older adults and 26.84 (SD = 2.03) among caregivers. Mean PHQ-9 scores were low in both groups, indicating minimal depressive symptoms at the group level.

Participants from the two museum sites differed descriptively in age distribution. Caregivers at Museum XXX were older on average than caregivers at the XXX Museum, while older adults at Museum XXX were also older on average than older adults at the XXX Museum. Because the main purpose of the study was to examine task-related alpha modulation during digital photo-supported storytelling rather than to compare museum sites, site-specific characteristics were reported descriptively.

### 3.2 gital photo-elicited storytelling participation

All dyads completed the museum-based digital photo-elicited storytelling procedure. Participants used mobile-phone photography during the museum visit to capture personally meaningful objects, scenes, or exhibition spaces. Each participant then selected one photograph to serve as a digital prompt for the storytelling interaction.

The selected photographs provided a shared visual reference for communication between older adults and caregivers. During the EEG-recorded storytelling task, dyad members alternated between speaking and listening roles. Older adults first narrated stories based on their selected photographs while caregivers listened, after which caregivers narrated their own selected photographs while older adults listened. This structure allowed the study to examine both storytelling and listening as active components of technology-supported social interaction.

### 3.3 Task-related alpha modulation during storytelling and listening

Task-related alpha modulation was examined using relative alpha power averaged across the predefined 10-electrode sensor-level set. Eyes-open resting was used as the within-participant baseline. Table 2 summarizes alpha power during eyes-closed resting, storytelling, and listening relative to eyes-open resting for older adults and caregivers.

In both participant roles, alpha power was significantly higher during eyes-closed resting than during eyes-open resting. Among older adults, relative alpha power increased from 12.04 (SD = 4.65) during eyes-open resting to 23.05 (SD = 7.27) during eyes-closed resting, Δ = 11.01, 95% CI [8.19, 13.84], t(31) = 7.96, p < .001, q < .001, dz = 1.41. Among caregivers, relative alpha power increased from 11.05 (SD = 4.87) during eyes-open resting to 30.16 (SD = 12.96) during eyes-closed resting, Δ = 19.11, 95% CI [14.64, 23.58], t(31) = 8.72, p < .001, q < .001, dz = 1.54. These findings supported the expected alpha increase during eyes-closed resting.

During digital photo-elicited storytelling, alpha power decreased significantly relative to eyes-open resting in both older adults and caregivers. Among older adults, alpha power decreased from 12.04 (SD = 4.65) during eyes-open resting to 7.77 (SD = 2.85) during storytelling, Δ = −4.27, 95% CI [−5.62, −2.91], t(31) = −6.41, p < .001, q < .001, dz = −1.13. Among caregivers, alpha power decreased from 11.05 (SD = 4.87) during eyes-open resting to 7.29 (SD = 2.17) during storytelling, Δ = −3.76, 95% CI [−5.37, −2.15], W = 41, p < .001, q < .001, r = −0.81.

Alpha power also decreased significantly during listening relative to eyes-open resting. Among older adults, alpha power decreased from 12.04 (SD = 4.65) during eyes-open resting to 7.80 (SD = 3.53) during listening, Δ = −4.24, 95% CI [−5.62, −2.85], t(31) = −6.24, p < .001, q < .001, dz = −1.10. Among caregivers, alpha power decreased from 11.05 (SD = 4.87) during eyes-open resting to 7.59 (SD = 3.19) during listening, Δ = −3.46, 95% CI [−4.67, −2.26], W = 15, p < .001, q < .001, r = −0.96.

These findings indicate robust alpha-band modulation across the digital photo-elicited storytelling task. Alpha activity increased during eyes-closed resting and decreased during both storytelling and listening. The decrease during listening suggests that listening in this digitally supported dyadic task was not a passive background condition but an active component of social-cognitive engagement.

### 3.4 Secondary cognitive and dyadic EEG findings

Secondary analyses examined whether central alpha activity was associated with cognitive performance and whether true dyads showed greater alpha-band inter-brain similarity than shuffled dyads. Associations between MoCA scores and Cz alpha power were weak and condition-specific. None of these associations remained significant after false discovery rate correction. These findings do not support a robust association between cognitive screening performance and central alpha activity in this sample.

Dyad-level alpha-band inter-brain similarity was also examined using spatial alpha-power patterns across the predefined 10-electrode set. During storytelling, observed dyad-level alpha similarity was modestly higher than within-site shuffled-dyad estimates. However, this effect did not remain significant after correction across conditions. Listening and resting conditions also did not show robust corrected effects. Therefore, dyad-level alpha similarity findings were interpreted as exploratory and hypothesis-generating rather than as evidence of reliable neural synchrony.

### 3.5 Summary of findings

Overall, the main finding was that digital photo-elicited storytelling and listening were associated with significant reductions in alpha power relative to eyes-open resting in both older adults and caregivers. These changes were observed alongside the expected increase in alpha power during eyes-closed resting. Secondary analyses of MoCA–Cz alpha associations and dyad-level alpha similarity were weak or non-robust after correction. The results therefore support the sensitivity of alpha-band EEG activity to technology-supported storytelling and listening, while indicating that cognitive and dyadic similarity findings should be interpreted cautiously.

## 4. Discussion

### 4.1 Principal findings

This study examined digital photo-elicited storytelling as a technology-supported social-cognitive activity in older adult–caregiver dyads. By combining mobile-phone photography, museum-based storytelling, and EEG recording, the study investigated how personally meaningful digital image prompts structured narrative interaction and how alpha-band activity changed during storytelling and listening.

Three main findings emerged. First, alpha power increased during eyes-closed resting and decreased during both storytelling and listening relative to eyes-open resting in older adults and caregivers. This pattern indicates robust task-related alpha modulation during digital photo-supported narrative interaction and is consistent with prior interpretations of alpha-band activity as sensitive to attentional control and task-related engagement (Klimesch, 2012; Pfurtscheller & Lopes da Silva, 1999). Second, associations between MoCA scores and Cz alpha power were weak, condition-specific, and did not survive false discovery rate correction. Third, dyad-level alpha-band inter-brain similarity during storytelling was modestly higher than within-site shuffled-dyad estimates, but this effect was not robust after correction across conditions. Together, the findings suggest that alpha-band activity was sensitive to participation in digital photo-elicited storytelling and listening, while cognition-related and dyad-level similarity findings should be interpreted cautiously.

### 4.2 Digital photo-elicited storytelling as technology-supported human behavior

The main contribution of this study is not simply that EEG changed during a storytelling task, but that a simple digital technology—mobile-phone photography—was used to structure meaningful human interaction. Digital photographs functioned as personally relevant prompts that allowed participants to select, organize, and communicate memories, meanings, and reflections. This is consistent with photo-elicitation approaches, in which photographs can evoke information, feelings, and memories that may not emerge through verbal questioning alone (Harper, 2002; Mysyuk & Huisman, 2020). In this sense, the technology did not replace human interaction; rather, it mediated and supported interaction by providing a shared visual reference for narrative exchange.

This framing is important for research on computers and human behavior because the behavioral value of digital technology often lies in how it shapes attention, memory, communication, and social participation. In the present study, mobile-phone photography enabled older adults and caregivers to transform museum experiences into individualized narrative material. Unlike standardized laboratory stimuli, the photographs were personally selected and emotionally meaningful. At the same time, the use of digital image prompts provided enough procedural structure to support systematic EEG recording and analysis.

The study therefore extends digital photo-elicitation beyond qualitative interviewing or reminiscence practice by linking it with psychophysiological measurement during live dyadic interaction. This approach may be useful for studying technology-supported activities in later life, particularly when the goal is to understand how accessible digital tools support meaningful communication rather than to evaluate complex or highly specialized technologies.

### 4.3 Storytelling and listening as active components of dyadic engagement

The findings also support the interpretation that both storytelling and listening are active components of digitally supported social interaction. Storytelling required participants to retrieve autobiographical or personally meaningful memories, organize narrative content, and communicate with a partner. Listening required sustained attention, interpretation of narrative meaning, emotional presence, and responsiveness to the speaker. Thus, listening should not be treated as a passive comparison condition.

This point is especially relevant for older adult–caregiver dyads. Caregivers are often conceptualized as facilitators, companions, or providers of assistance, but in narrative interaction they also serve as listeners, responders, and co-constructors of meaning. Digital photo-elicited storytelling may therefore provide a practical setting in which caregivers and older adults can engage in mutual reflection. The EEG findings cannot be interpreted as direct evidence of empathy, but they do indicate that the listening condition was associated with task-related alpha modulation rather than a resting-like pattern.

For CHB readers, this suggests that the behavioral effects of digital tools should be understood relationally. A photograph does not only prompt an individual memory; it can also organize shared attention, conversational turn-taking, and interpersonal meaning-making. This view is consistent with broader aging research showing that socially integrated and cognitively active lifestyles are important for later-life cognitive health and participation (Fratiglioni et al., 2004; Livingston et al., 2024). Future studies should therefore combine digital interaction paradigms with behavioral coding of speech, gaze, silence, turn-taking, and listener responses to clarify how technology-supported storytelling unfolds between partners.

### 4.4 Alpha-band modulation during digital photo-supported storytelling

Alpha power decreased during storytelling and listening relative to eyes-open resting in both older adults and caregivers. This pattern is consistent with the interpretation that alpha-band activity is sensitive to attentional engagement, controlled access to stored information, and task-related information processing (Klimesch, 2012). It is also consistent with the broader event-related desynchronization framework, in which decreases in alpha or beta power are interpreted as reflecting activation of task-relevant neural systems during cognitive or sensorimotor processing (Pfurtscheller & Lopes da Silva, 1999).

Storytelling and listening required participants to engage with personally meaningful content, maintain attention, process narrative information, and participate in dyadic communication. Alpha suppression during these conditions may therefore reflect a shift from resting-state activity toward active cognitive-social engagement. This interpretation is also compatible with evidence that oscillatory mechanisms contribute to memory encoding and retrieval, including neocortical alpha/beta dynamics and hippocampal theta-related mechanisms (Griffiths et al., 2021; Herweg et al., 2020; Karlsson & Sander, 2023).

The finding that alpha decreased during both speaking and listening is important. It suggests that the digitally supported task engaged both members of the dyad across communicative roles. Although the roles differed behaviorally, both required active participation. This supports the idea that digital photo-elicited storytelling can be used as a structured but naturalistic paradigm for examining cognitive and social engagement in later life.

However, the present EEG findings should be interpreted at the sensor level. The study did not use source localization, and individual alpha peak frequency was not controlled. Therefore, alpha modulation should not be treated as evidence of localized neural mechanisms or as a diagnostic marker. Instead, alpha power is best interpreted as a supplementary physiological indicator of task-related engagement during digital photo-supported interaction.

### 4.5 Cognitive and dyadic EEG findings

The secondary MoCA–Cz alpha findings were weak and did not survive correction for multiple comparisons. This suggests that central alpha activity during the task should not be interpreted as a robust marker of cognitive screening performance in this sample. One possible explanation is that digital photo-elicited storytelling involves multiple processes, including memory retrieval, attention, emotion, language, social communication, and partner responsiveness. A single central alpha measure may be too limited to capture this complexity.

The dyadic inter-brain similarity findings were also exploratory. True dyads showed modestly higher alpha-band spatial similarity during storytelling than within-site shuffled dyads, but this effect did not remain significant after correction across conditions. This result should not be interpreted as definitive evidence of neural synchrony. Prior hyperscanning studies have shown that inter-brain measures can be associated with social interaction and naturalistic interpersonal engagement (Dumas et al., 2010; Kinreich et al., 2017). However, similarity between dyad members may also reflect shared visual context, common task structure, museum environment, or role-based timing, in addition to any dyad-specific interpersonal coordination.

Nevertheless, the surrogate-dyad approach is useful because it provides a more cautious comparison than interpreting raw dyadic similarity alone. For future CHB-oriented research, dyadic EEG measures should be paired with behavioral data from the interaction itself, such as speech timing, mutual gaze, conversational alignment, emotional tone, and caregiver response patterns. These combined measures would help determine whether physiological similarity is meaningfully related to technology-supported social behavior.

### 4.6 Implications for technology-supported aging and caregiving

The findings have several implications for research and practice. First, they suggest that simple and familiar digital tools can support meaningful social-cognitive activity in older adult–caregiver dyads. Mobile-phone photography is accessible, low-cost, and already familiar to many participants, making it a practical medium for memory sharing and communication.

Second, the study highlights the importance of designing digital activities that support relational participation rather than individual performance alone. In the present protocol, the digital photograph served as a shared object of attention. This allowed both older adults and caregivers to participate in storytelling and listening, and it positioned caregivers as active interaction partners rather than background supporters.

Third, the study demonstrates the feasibility of combining technology-supported interaction with psychophysiological measurement in real-world settings. EEG should not replace qualitative, behavioral, or relational assessment, but it can provide supplementary evidence about physiological engagement during meaningful activity. This may be particularly valuable for studying older adults and caregivers in ecologically valid contexts where traditional laboratory tasks may not capture the richness of everyday communication.

### 4.7 Limitations and future directions

Several limitations should be considered. First, the sample size was modest, particularly for dyad-level analyses. The study was better suited to detecting moderate task-related effects than small cognitive or interpersonal effects. Second, participants were recruited from museum-based activities, which may limit generalizability. Individuals who attend museum programs may be more socially engaged, mobile, or comfortable with cultural participation than the broader older adult and caregiver population.

Third, the study did not include a non-digital storytelling condition, a passive viewing condition, or a non-museum control condition. Therefore, the findings cannot isolate the specific contribution of mobile-phone photography from the broader effects of museum context, storytelling, social interaction, or personally meaningful content. Fourth, listening quality was inferred from the task context and was not directly measured using behavioral coding or an empathy scale. Therefore, the study should not be interpreted as directly measuring empathy.

Fifth, EEG analyses were conducted at the sensor level using relative alpha power. The findings should not be interpreted as evidence of localized neural generators, clinical diagnosis, or definitive neural mechanisms. Sixth, dyad-level inter-brain similarity was based on spatial alpha-power similarity rather than continuous time-resolved hyperscanning connectivity. Future studies should use larger samples, preregistered hypotheses, behavioral coding, time-resolved dyadic EEG methods, and longitudinal designs to examine how digital photo-elicited storytelling affects memory, communication, caregiver engagement, and relationship quality over time.

## 5. Conclusion

This study suggests that digital photo-elicited storytelling can provide a meaningful and feasible medium for studying technology-supported social-cognitive engagement in older adult–caregiver dyads. Alpha power decreased during both storytelling and listening relative to eyes-open resting, indicating task-related modulation during digitally supported narrative interaction. However, associations between central alpha activity and cognitive performance were weak, and dyad-level alpha similarity findings were exploratory and non-robust after correction.

The study contributes to Computers in Human Behavior by showing how a simple digital tool, mobile-phone photography, can structure memory sharing, communication, and dyadic engagement in later life. EEG offered supplementary physiological evidence of engagement, but the core behavioral contribution lies in understanding how digital image prompts support meaningful human interaction between older adults and caregivers.

## Data Availability

All data produced in the present study are available upon reasonable request to the authors.

