## Supplementary material for "Digital Photo-Elicited Storytelling and Alpha-Band EEG Dynamics in Older Adult–Caregiver Dyads": Table1-2

**Table 1**

*Participant characteristics by role and museum site.*

| Group | n | Age, years | PHQ-9 | MoCA | Female, n (%) | Male, n (%) |
|---|---|---|---|---|---|---|
| Total sample | 64 | 56.70 ± 14.92 | 1.69 ± 2.20 | 25.69 ± 3.33 | 49 (76.6%) | 15 (23.4%) |
| Caregivers, total | 32 | 44.03 ± 9.43 | 2.16 ± 2.76 | 26.84 ± 2.03 | 26 (81.2%) | 6 (18.8%) |
| Older adults, total | 32 | 69.38 ± 5.68 | 1.22 ± 1.31 | 24.53 ± 3.95 | 23 (71.9%) | 9 (28.1%) |
| Caregivers – Museum AAA | 16 | 49.62 ± 5.16 | 1.50 ± 1.10 | 27.50 ± 1.71 | 15 (93.8%) | 1 (6.2%) |
| Older adults – Museum AAA | 16 | 72.31 ± 6.24 | 1.38 ± 1.36 | 24.38 ± 3.96 | 12 (75.0%) | 4 (25.0%) |
| Caregivers – BBB Museum | 16 | 38.44 ± 9.51 | 2.81 ± 3.69 | 26.19 ± 2.17 | 11 (68.8%) | 5 (31.2%) |
| Older adults – BBB Museum | 16 | 66.44 ± 3.05 | 1.06 ± 1.29 | 24.69 ± 4.06 | 11 (68.8%) | 5 (31.2%) |

*Note:* Values are mean ± standard deviation unless otherwise stated. PHQ-9 = Patient Health Questionnaire-9; MoCA = Montreal Cognitive Assessment. Museum AAA and BBB Museum each contributed 16 dyads.

**Table 2**

*Task-related alpha modulation during digital photo-elicited storytelling and listening.*

| Role | Comparison | Baseline EO, M (SD) | Condition M (SD) | Δ Mean [95% CI] | Test statistic | p | FDR q | Effect size |
|---|---|---|---|---|---|---|---|---|
| Older adults | EC vs EO | 12.04 (4.65) | 23.05 (7.27) | 11.01 [8.19,13.84] | t(31) = 7.96 | < .001 | < .001 | dz = 1.41 |
| Older adults | Storytelling vs EO | 12.04 (4.65) | 7.77 (2.85) | −4.27 [−5.62,−2.91] | t(31) = −6.41 | < .001 | < .001 | dz = −1.13 |
| Older adults | Listening vs EO | 12.04 (4.65) | 7.80 (3.53) | −4.24 [−5.62, −2.85] | t(31) = −6.24 | < .001 | < .001 | dz = −1.10 |
| Caregivers | EC vs EO | 11.05 (4.87) | 30.16 (12.96) | 19.11 [14.64, 23.58] | t(31) = 8.72 | < .001 | < .001 | dz = 1.54 |
| Caregivers | Storytelling vs EO | 11.05 (4.87) | 7.29 (2.17) | −3.76 [−5.37, −2.15] | W = 41 | < .001 | < .001 | r = −0.81 |
| Caregivers | Listening vs EO | 11.05 (4.87) | 7.59 (3.19) | −3.46 [−4.67, −2.26] | W = 15 | < .001 | < .001 | r = −0.96 |

Note. EO = eyes-open resting baseline; EC = eyes-closed resting. Values are relative alpha power averaged across the predefined 10-electrode sensor-level set: Fp1, Fp2, F3, F4, F8, T3, T4, Fz, Cz, and Pz. For normally distributed difference scores, paired-samples t-tests were used and effect sizes are reported as Cohen's dz. For non-normal difference scores, Wilcoxon signed-rank tests were used and effect sizes are reported as rank-based r. FDR q values were calculated using the Benjamini–Hochberg procedure.
